# The impact on clinical success from the 23andMe cohort

**DOI:** 10.1101/2024.06.17.24309059

**Authors:** Xin Wang, Sotiris Karagounis, Suyash S. Shringarpure, Rohith Srivas, Qiaojuan Jane Su, 23andMe Research Team, Vladimir Vacic, Steven J. Pitts, Adam Auton

## Abstract

90% of therapeutic programs that enter clinical trials ultimately fail. Human genetic variation provides a set of “natural experiments” that can inform successful strategies for therapeutic discovery. Previous work has estimated that drug targets with human genetics supported mechanisms have a 2-3x increased likelihood of succeeding in the clinic compared to those without. 23andMe, Inc. is a direct-to-consumer genetics company that has created a human genetics dataset approximately an order of magnitude larger in sample size than current publically available cohorts. As of 2024, 23andMe has approximately 15 million individuals with genotype and phenotype data, of which ∼80% consent to participation in research. In this work, we explore how both the scale of the genetic data and improved methods to link genetic associations to putative causal genes impact the prediction of clinical success. Comparing the total number of target-indication pairs that have reached at least phase I that are also supported by genetic evidence, the number of target-indication pairs with support from 23andMe is 60% greater than that with support from all GWAS datasets in the public domain. Including 23andMe genetic evidence approximately doubles the number of target-indication pairs in the clinic that are supported by human genetics. Furthermore, we show that genetic associations derived from entirely self-reported phenotypes are 2-3x enriched for clinical success, just as for clinically derived phenotypes. In contrast to conclusions from the recent publication of Minikel *et al.*, we found that minor allele frequencies and effect sizes from GWAS influence the relative success estimates for program approvals, and that drug programs supported by rare and large effect associations have greater (3-4x) likelihood to be approved compared to common variant associations with small effects. Finally, improved gene mapping to identify the likely causal genes underlying genetic associations can result in up to 4-5x enrichment for trial success. With the increased power and scale of the 23andMe genetic dataset, we identify an expansive set of opportunities that may be pursued in the clinic, emphasizing the importance of cohort size and gene mapping confidence in deriving clinical value.

## Introduction

Human genetic variation provides a set of “natural experiments” that can inform successful strategies for therapeutic discovery^1^. In recent years, availability of large-scale human genetics datasets has increased via the creation of biobanks and cohorts such as the UK Biobank^2^, All of Us^3^, FinnGen^4^, etc. These datasets can range from several hundred thousand to approximately 1 million individuals in cohort size, and have provided substantial value in unifying large-scale genotyping with deeply phenotyped datasets^5^. 23andMe, Inc. is a direct-to-consumer genetics company that has created a human genetics dataset approximately an order of magnitude larger in sample size than the current publically available cohorts. At the time of writing, over 12 million 23andMe customers with genotype and phenotype data have consented to participate in research. Previous studies have estimated that drug targets with human genetic evidence are 2-3 times more likely to achieve clinical success compared to those without^6–8^. However, these studies have not fully addressed questions regarding the scale of the underlying genetic evidence, strategies of phenotype and genotype ascertainment, or confidence in gene hypotheses derived from genetic associations and their impact on clinical success.

Self-report data are commonly used within the field of genetics. Web or smart phone-based questionnaires are highly efficient for collecting large amounts of phenotype data in human genetics studies, and can be deployed at the scale necessary to power genome-wide association studies (GWAS). The UK Biobank derives phenotypes using electronic medical records (EMRs), questionnaire and interview responses, physical measurements, and family history^9^. The All of Us research program utilizes EMRs in combination with physical measurements, survey responses, and data from wearables^10,11^. 23andMe collects phenotype data primarily through self-reported survey responses. Early studies demonstrated that genetic associations discovered utilizing self-reported medical data replicated those derived from clinical sources at the expected rate^12^. More recently, it was shown that GWAS using phenotypes defined using hospital records versus questionnaires find largely similar genetic effects genome-wide^13^. Nonetheless, doubts remain that self-reported data can be of value in a target discovery context due to the possibilities of mis-reporting disease diagnosis both in terms of individuals reporting disease diagnoses when no such diagnosis exists and individuals failing to report a disease diagnosis, which could plausibly lead to loss of statistical power or false discoveries in subsequent GWAS analyses^13–16^.

Among the variations we observe in the genome, variants that have protein coding consequences can be particularly valuable for interpreting genetic associations. Due to the large number of rare variants in the human genome, and that variants with functional consequences tend to be rare, current studies remain under-powered. In recent years, there has been an increasing amount of available whole exome and whole genome sequencing datasets, leading to a number of exciting findings derived from hundreds of thousands of exomes^17–19^. However, sequencing hundreds of thousands of individuals remains very costly and prohibitive compared to array based genotyping. With new strategies to impute rare variants^20^, it is important to evaluate the cost-benefit of sequencing based association studies and those that leverage genotype imputation.

Even after genetic analyses have been performed, not all genetic evidence is of equal value for target discovery purposes. For example, in the GWAS framework, the genetic associations usually do not directly pinpoint a causal gene. Identifying the specific causal variants for a given genetic association can be challenging due to many variants being in high levels of linkage disequilibrium (LD). Even if the causal variant can be identified with confidence, there may remain significant ambiguity regarding the functional impact of the causal variant, and uncertainty regarding which gene(s) is being modulated. This uncertainty ensures that not all genetic evidence can be considered equal for downstream target discovery activities.

In this work, we performed a refined analysis of drug target success using data from 23andMe to address the above questions. We show that with the greater scale, 23andMe data provides genetic support for a greater number of target-indication(T-I) pairs in the clinic compared to the total aggregate of all publicly available data, and in combination with public data, doubles the coverage on total T-I pairs that are supported by human genetics. We further show that drug programs supported by genetic associations derived from entirely self-reported phenotypes are 2-3 times enriched for success, and that the increased statistical power derived from the larger scale and improved gene mapping of 23andMe can result in up to 4-5 times enrichment for clinical approvals, emphasizing the importance of cohort size and gene mapping confidence in deriving clinical value.

## Methods

### 23andMe phenotypes

23andMe phenotypes were derived from survey responses provided by 23andMe research participants, and mapped to the Experimental Factor Ontology (EFO)^21^ using algorithmically mapped EFO terms followed by manual curation (Supplementary Table 1). Automatic mapping was done via text2term ontology mapping^22^ and via strictly matching ontology terms by matching standardized tokens of input text with standardized tokens of EFO labels or aliases. To select phenotypes of therapeutic relevance, we identified all EFO terms associated with an indication listed in Citeline Pharmaprojects and selected 23andMe phenotypes that mapped to one of these EFO terms with similarity > 0.9. We augmented this with a manually curated set of disease phenotypes from public sources (i.e. from OpenTargets, Genebass, intOGen, OMIM, and PICCOLO)^23^, which we also mapped to a 23andMe phenotype with EFO similarity > 0.9. The EFO similarity scores are computed based on the relative information contents between the least common ancestor node and the individual nodes in the ontology (Lin’s method^24^). The sensitivity of relative success analysis to the EFO threshold used was assessed (Supplementary Figure 1), and a threshold of 0.9 is used throughout the rest of this work.

### GWAS and conditional analyses

A total of ∼7.5 million consented individuals were included in the 23andMe dataset for GWAS analyses. Participants provided informed consent and volunteered to participate in the research online, under a protocol approved by the external AAHRPP-accredited IRB, Ethical & Independent (E&I) Review Services. As of 2022, E&I Review Services is part of Salus IRB (https://www.versiticlinicaltrials.org/salusirb). GWAS was conducted separately for each genetic ancestry group classified as African American, East Asian, European, Latino, or South Asian based on inferred local ancestry assignments from 23andMe^25^. For each phenotype, a maximal set of unrelated individuals were identified using a segmental identity-by-descent (IBD) algorithm^26^. Two individuals were considered unrelated if they shared less than 700cM IBD. A linear or logistic regression model was fit for quantitative and case-control phenotypes respectively, taking into account age, sex, genetic principal components (from five to nine depending on genetic ancestry group), and genotyping platforms. Association was performed with both genotyped and imputed variants. UK Biobank whole genome sequence data^27^ that are publicly available combined with 23andMe internal sequencing data, a total of over 200,000 whole genome sequences, were used for imputation, following phasing of microarray genotype data. There were over 300 million imputed variants, and over 172 million high quality variants after quality control. Association testing p-values were derived from a likelihood ratio test. LD-score regression (LDSC)^28^ intercepts were used to correct for inflation in European GWAS, and genomic control (GC)^29^ was used for other genetic ancestry groups. Genome-wide significant associations (p-value < 5e-8) were identified and grouped into “loci”, defined as all variants with p-value < 1e-6 and that no two loci have distance smaller than 250kb.

Following the marginal association procedure, a forward stepwise regression was performed in order to identify additional independent associations within a locus. The GWAS marginal index variant was added to the regression model, and for each following step, the variant with the smallest p-value was further added to the regression model. The stepwise regression concludes if the most significant variant of a step had p-value > 1e-5, or when a total of 20 steps have been reached.

For lead variants identified in the conditional stepwise procedure, an additional regression was performed by leaving them out one at a time and recomputing the association test statistics for all variants in the locus using the remaining lead variants as covariates. The variant with the smallest p-value in each regression was re-selected as the lead variant.

### Burden tests

Associations between imputed rare variants and a phenotype was conducted via a collapsing burden test framework for each gene^30^. Cumulative burden for a gene was derived as the probability of each haplotype containing at least one qualifying variant and taking the sum from both haplotypes. Specifically, for individual sample *i*, the gene burden *S_i_* was calculated as

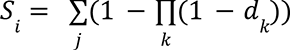

for each haplotype *j* where *d* represents the imputed dosage of each qualifying variant *k* in the gene. Gene burdens were computed for variants with minor allele frequency (MAF) < 1% and predicted as 1) high confidence (HC) loss-of-function as predicted by LOFTEE (v1.0.4)^31^ or 2) HC loss-of-function from LOFTEE or missense variants with gMVP^32^ score greater than 0.8. All predictions were based on the canonical transcript (GRCh38, Gencode V43). A linear (for quantitative traits) or penalized logistic (for binary traits) regression was performed using the burden score as genotype dosages and correcting for age, sex, and five genetic principal components. Final burden results were corrected for nearby independent common GWAS associations using COJO^33^.

### Gene mapping

Various variant to gene (V2G) approaches were used to identify putative causal genes for each GWAS association in the 23andMe dataset. First, associations from GWAS were mapped to likely causal genes via coding variants (stop-gain, start/stop lost, frameshift, missense, inframe insertion/deletion, and splice site changes) identified as being in high LD with the GWAS index variant (r^2^ >= 0.8). Second, putative causal genes were found by identifying expression, protein, and splice QTLs that were in high LD with the GWAS index variant (r^2^ >= 0.8). Third, colocalization^34^ was performed between the GWAS and QTL summary statistics where available, and a putative causal gene was identified when the posterior probability of having a shared common variant (H4) >= 0.8. Finally, the nearest gene to a given GWAS association, as measured by distance from an index variant to the nearest transcription start site of a canonical transcript, was also used as a gene mapping method. Loci where there were greater than three gene hypotheses were removed. When several coding variants or eQTL in high LD (r^2^ >= 0.8) with independent GWAS or conditional analysis index variant supported the same gene hypothesis, we defined a gene hypothesis as having an “allelic series”. Non-23andMe GWAS data used gene mapping methods as described previously^23^.

### Power calculation

For a case-control phenotype with total sample size *N* and prevalence (proportion of cases) *φ*, with respect to a variant with minor allele frequency *f*, and observed effect size on the log-odds scale for the minor allele being β, the non-centrality parameter of the χ2 distribution

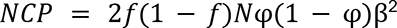

Taking into account attenuation due to imputation (quality R^2^), the non-centrality parameter becomes

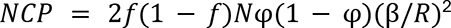

Power is calculated as the probability of rejecting the null hypothesis that there is no association of the phenotype with the variant at genome-wide significance threshold (p-value = 5e-8) with 1 degree of freedom.

### V2G weighting and V2G score

In order to provide a quantitative measure of the strength of V2G evidence, we created a score that combined multiple levels of V2G evidence. Gene mapping evidence within a locus was categorized into distinct features that were used to train a model that predicted the set of genes known to be causal in a gold- or silver-standard dataset. For each gene, the features used in the model were: number of coding (putative loss-of-function or missense) variants in high LD (r^2^ > 0.8) with a GWAS index variant, number of LoF variants in high LD with a GWAS index variant, number of independent QTL datasets with at least one QTL (expression QTL, protein QTL, splice QTL p-value <= 1e-7) in high LD with a GWAS index variant, number of genes in a locus (500kbp surrounding the GWAS index variant), and whether a gene is the weighted nearest gene as measured by the distance from variants in the 95% credible set to the transcription start site of a gene in the locus weighted by their posterior inclusion probability (PIP)^35,36^. Only variants with PIP >= 0.001 were used.

High confidence gene-phenotype pairs in the gold-standard set curated by Open Targets^36^, as well as an evaluation dataset leveraging fine-mapping results from 95 traits in the UK Biobank proposed previously^37^ were used as “truth” sets in training. 23andMe associations were assigned to “true” causal genes based on the above-mentioned truth set by matching phenotype and genes, resulting in 368 loci. A nested cross-validation framework (Supplementary Figure 4) was used in training a stochastic gradient boosting model. Five outer folds and five inner folds were used, where chromosomes were used in determining the outer folds such that each chromosome only belonged to a single cross-validation fold as the held-out testing set. For each outer fold, five inner fold cross-validation were performed to tune model parameters.

A V2G score was assigned to each gene based on the final model. For each phenotype, the model assigned gene scores for each independent association, for example, within the same locus, marginal association and conditional association(s) would be considered independently. The final score for a gene is the summation of all scores for the gene across all relevant independent association signals.

## Results

### Target-indications supported by self-reported data are 2-3 times enriched for clinical success

To compare therapeutically relevant phenotypes from the 23andMe database to clinical development programs, we used all EFO terms associated with an indication listed in Citeline Pharmaprojects and mapped it to a 23andMe phenotype with similarity > 0.9 (see Methods). A total of 110,249 gene-phenotype pairs across 13,552 genes (693 GWAS phenotypes) were identified, including gene hypotheses derived from statistically independent associations identified by conditional fine-mapping^34^ using any of the V2G methods as described (see Methods). Only associations with p-value <= 5e-8 were included in the analysis. Leveraging previously curated Pharmaprojects indications^23^, we considered indications with a matched GWAS phenotype^23^ that had at least 1 genome-wide significant association as having any genetic support for target hypotheses from GWAS. Of the 1,033 unique indications identified from Pharmaprojects, 745 (72%) had genetic support from 23andMe data, compared to 667 (65%) found in the public domain, a 12% relative increase in coverage.

We used relative success (RS) as previously defined^23^ to directly compare human genetics evidence from 23andMe to that obtained from the public domain. We found that genetically supported T-I pairs using only 23andMe data had RS=2.17 [1.90, 2.47], which is comparable to the overall RS (2.28 [1.94, 2.67]) from Open Target Genetics (OTG) and all GWAS datasets considered in the public domain (RS=2.24 [1.92, 2.62]) (Figure 1A, p-value =0.74, test for proportions). Comparing the number of T-I pairs in Pharmaprojects that have reached at least phase I and are supported by genetic evidence, the total number of pairs with genetic support from 23andMe (1,021) is greater than that with support from all considered GWAS datasets in the public domain (632). Including 23andMe genetic evidence approximately doubles the number of T-I pairs in Pharmaprojects that are supported by human genetic evidence (1,443 versus 779) (Figure 1A, Supplementary figure 2), consistent with the observed trend that the number of associations discovered linearly increases with GWAS effective sample size for many disease phenotypes^16^ and demonstrating the power of discovery in this very large cohort.

**Figure 1.**
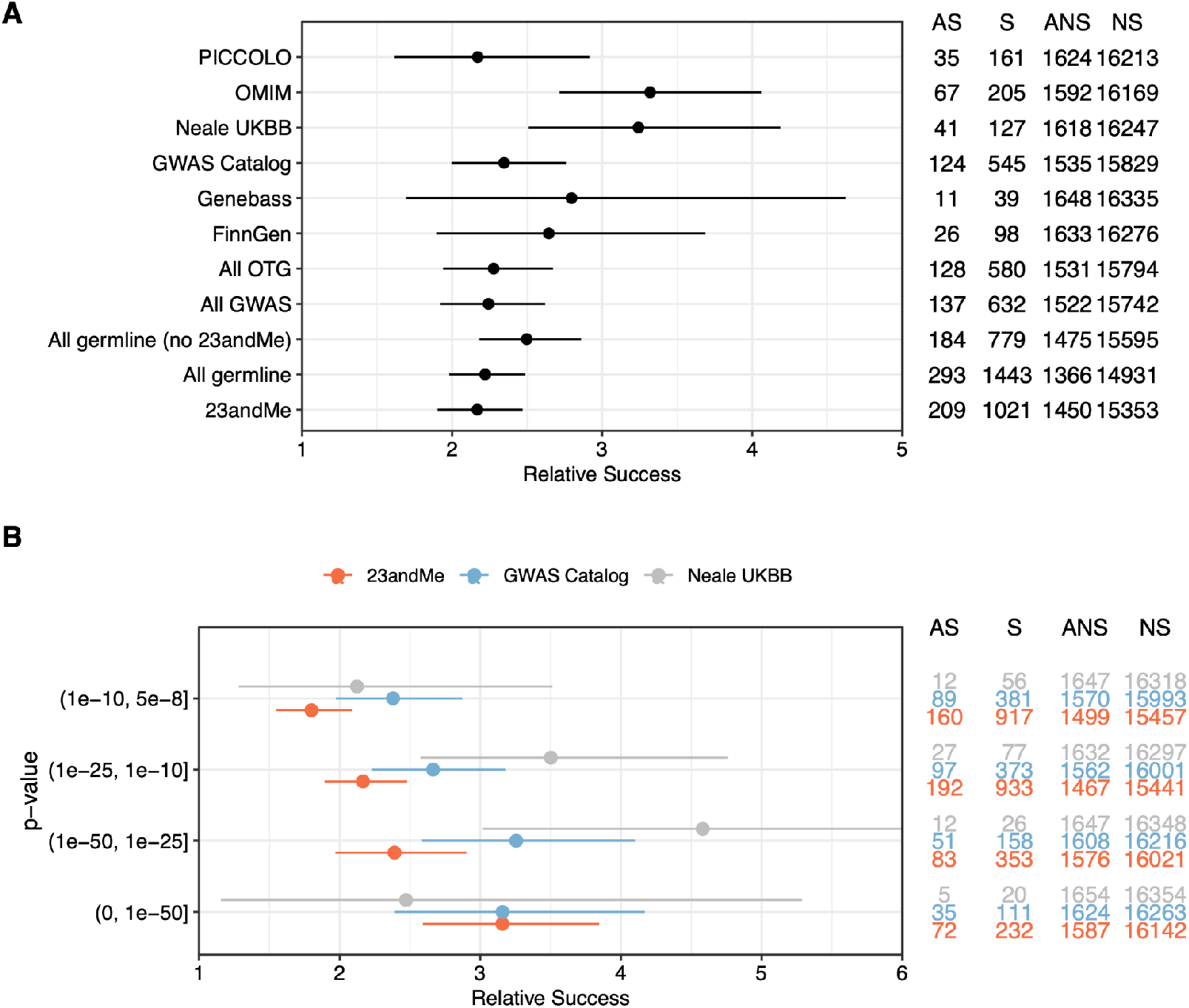
Target-indication pairs supported by 23andMe genetic evidence are 2-3 times enriched for clinical success. A. Relative clinical success (phase I to launch) comparing 23andMe and other human genetic evidence sources (GWAS p-value <= 1e-10, gene mapping described in Methods); B. Relative success in 23andMe data stratified by p-value in comparison to the GWAS Catalog and the UK Biobank. AS: T-I pairs approved and supported by genetic evidence; S: total T-I pairs supported by genetic evidence; ANS: T-I pairs approved but not supported by genetic evidence; NS: total T-I pairs not supported by genetic evidence. RS = (AS/S) / (ANS/NS).

As GWAS power improves, we expect to find more associations with smaller effect sizes or rarer allele frequencies, many also in the form of secondary / conditional associations. We looked into the relationship between the p-values of associations and their predictive value for relative success in clinical approvals, comparing data from 23andMe and those from the UK Biobank or the GWAS Catalog. We find a monotonic trend where smaller p-values are associated with greater clinical success in 23andMe data and the GWAS Catalog (Figure 1B, p-value=3.9e-6, Chi-squared test for trend). When focusing on GWAS p-values <= 1e-25, approximately a quarter (83/353) of T-I pairs are supported by genetic evidence from 23andMe, and increasing to approximately one-third (72/232) when focusing on p-values <= 1e-50.

### Rare large effect gene-indication associations are 3-4 times enriched for clinical success

Many variants that are likely to have important functional consequences, such as loss-of-function variants and deleterious missense variants, are often observed at lower frequencies in the general population. As such, the power to detect rare variants of even moderately large effects can be limited by the available sample size. As the scale of genetic databases has increased, we are now well positioned to explore the importance of rare variants (e.g. MAF <= 0.1%) for target discovery. The 23andMe cohort leverages internal and other available sequencing datasets (see Methods), allowing imputation down to frequencies of 1e-5 in Europeans and 1e-4 in a number of non-European populations (imputation aggregate R^2^ > 0.3).

Using the 23andMe database, we looked into how allele frequency and effect size influence relative success in drug approvals. Contrary to conclusions from previous work^23^, we found a statistically significant trend between MAF and RS (p-value = 1.0e-6, Chi-squared test for trend), and between odds ratio (OR) and RS (p-value = 0.0013, Chi-squared test for trend). GWAS associations where the index variant has MAF < 0.1% or has OR > 2 were ∼4 times enriched for clinical success (Figure 2A, B). The results remain significant if T-I pairs from OMIM were removed (Supplementary figure 3). This trend was not readily identifiable from the GWAS Catalog or the UK Biobank for several possible reasons: 1) individual GWAS sample sizes were smaller (median sample size ∼5,000 for GWAS catalog, median sample size ∼53,000 for UK Biobank); 2) rare variants were removed from association testing and as a result may not exist in the summary statistics; 3) imputation that tapped into rare variants, especially those that have MAF < 0.1% may be limited.

**Figure 2.**
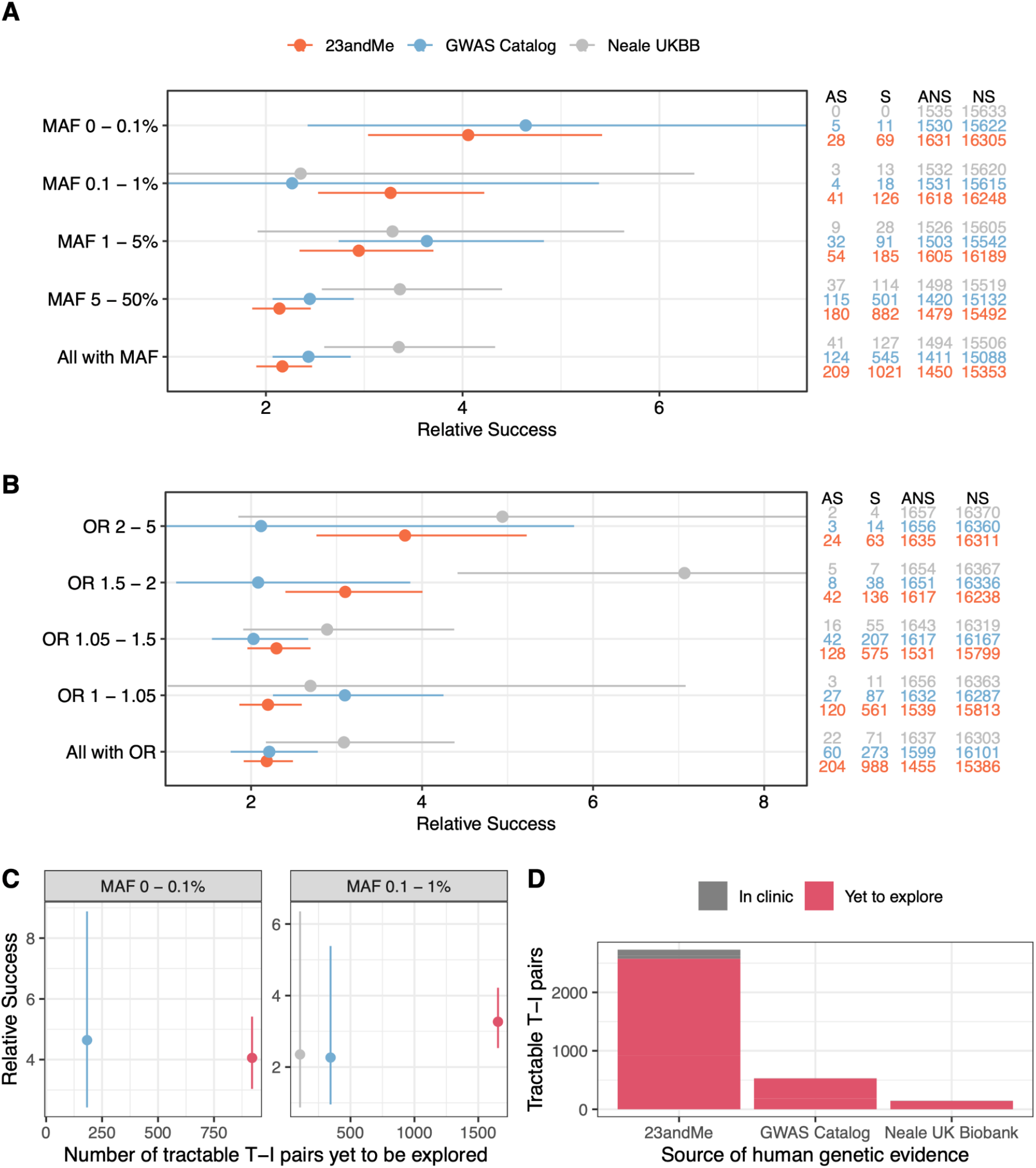
Rare, large effect genetic associations are 3-4 times enriched for clinical success. A-B. Relative success in 23andMe data stratified by MAF and OR in comparison to the GWAS Catalog and the UK Biobank; C. The number of tractable T-I pairs yet to be explored, stratified by MAF among variants with MAF < 1%; D. Comparison of tractable T-I pairs already in clinic versus those yet to be explored by source of human genetic evidence (MAF < 1%).

In recent years, with the efforts to generate whole exome and whole genome sequences in large population cohorts, particularly from the UK Biobank and followed by FinnGen, it is increasingly important to evaluate the cost-benefit of sequence-based analyses compared to imputation-based analyses. We took the rare associations (MAF < 1%) from 23andMe that provided genetic support to Pharmaprojects T-I pairs and conducted power analyses with respect to these associations relative to cohort size (Figure 3). As estimated previously, with a reference panel of 200,000 or more whole genome sequences, variants with MAF > 1% can be imputed at almost perfect accuracy, with variants that have minor allele count (MAC) of 5 having imputation accuracy (average imputation R^2^) at approximately 0.4^17^. As 23andMe includes rare variants with imputation aggregated R^2^ > 0.3, and that the 23andMe dataset used here has ∼7.5 million individuals, we therefore compared the power for finding associations varying total sequencing sample sizes from 50,000 to 2.5 million in order to take into account the attenuation in association testing due to imputation^38,39^, and assuming no attenuation in sequencing based association tests. We used discovery effect sizes in this calculation that could be biased by “winner’s curse”, hence this analysis may be optimistic, as the true effect sizes may be smaller than observed. For highly prevalent diseases (prevalence=20%, 50%), a cohort size of 500,000 is reasonably powered to discover these associations. However, at much lower prevalence (prevalence=1%, 4%), the majority of these rare associations have limited power to be detected in a sequencing cohort that would currently be widely available (Figure 3A). We also estimated the required sequencing cohort size in order to have 80% power to detect these rare variant associations found in 23andMe at genome-wide significance levels. We found that approximately 1 million sequenced individuals are required to find ∼50% of these associations, and a 3 million individual cohort is required in order to discover ∼95% of these rare associations (Figure 3B).

**Figure 3.**
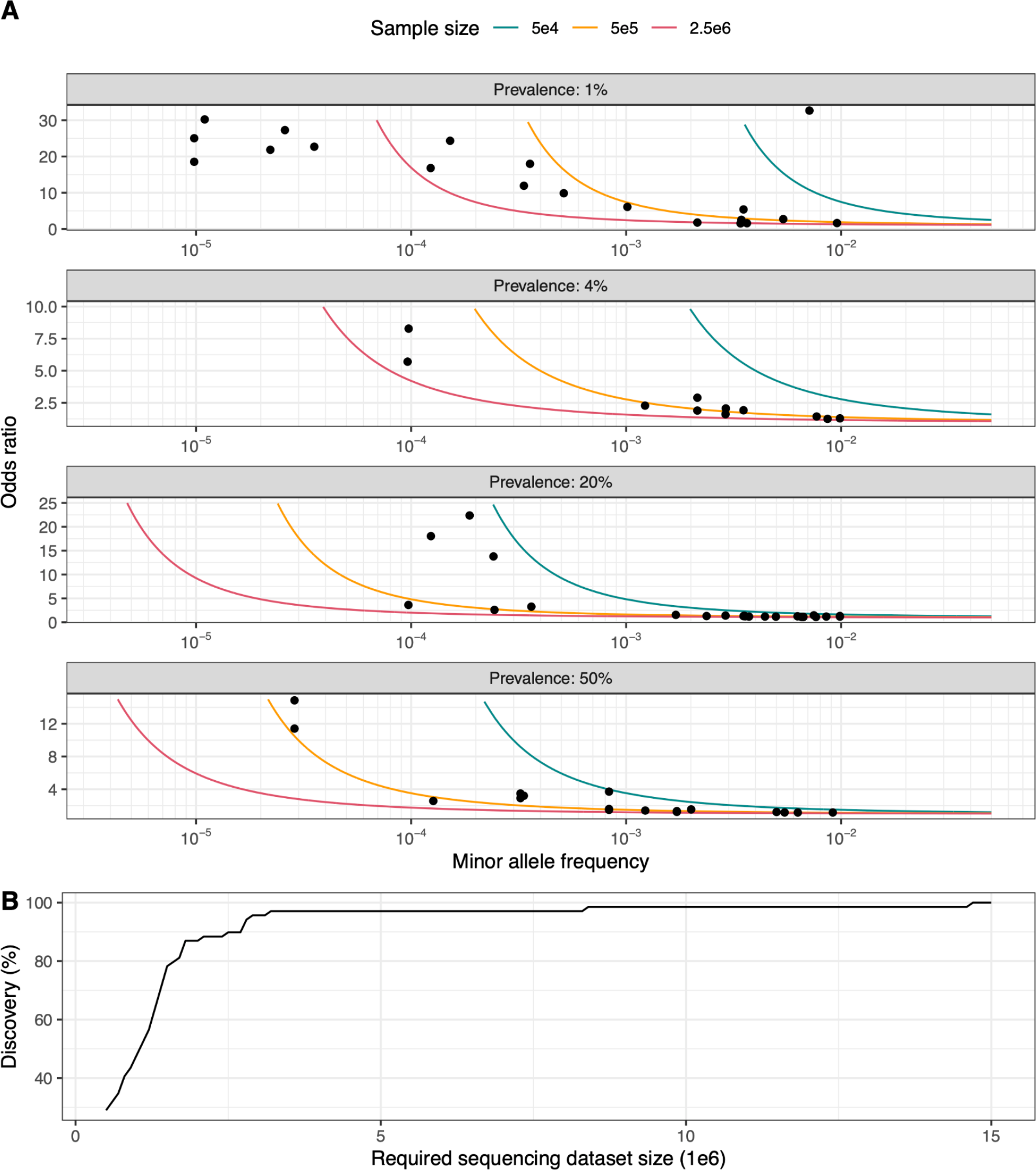
Power to identify rare variant associations that support Pharmaprojects target-indication pairs. A. GWAS power (see Methods) for different sample sizes (colored lines) stratified by quartiles of disease prevalence from 23andMe (panels). Points are associations with MAF < 1% from 23andMe that provided genetic support for approved T-I pairs in Pharmaprojects. For each panel, only associations from disease phenotypes with prevalence falling within the specified quartile are shown, i.e. (0,1%], (1%,4%], (4%,20%], (20%,50%] observed prevalence in each of the panels. Lines identify 80% power at the genome-wide significant p-value threshold of 5e-8; B. Percent discovery (y-axis) relative to required sequencing dataset sizes (x-axis) at 80% power and p-value threshold of 5e-8 for associations with MAF < 1% from 23andMe that provided genetic support for approved T-I pairs in Pharmaprojects. Minor allele frequencies, prevalences, effect sizes, as well as imputation average R^2^ are those obtained from 23andMe. Percent discovery is defined as the percentage of associations that have at least 80% power.

### Gene prioritization algorithm enriches for clinical success

Despite more associations being found with greater GWAS power, it remains a mountainous task to find the likely causal gene(s) that underlie an association. We further stratified the gene-phenotype pairs by V2G criteria and evaluated their influence on relative success. Coding variants, allelic series, and burden V2G result in approximately 3-fold relative success (Figure 4A), similar to that from OMIM supported T-I pairs (Figure 1A). Distance-based methods or eQTL provide weaker evidence with an approximately 2-fold relative success for clinical approval (Figure 4A).

**Figure 4.**
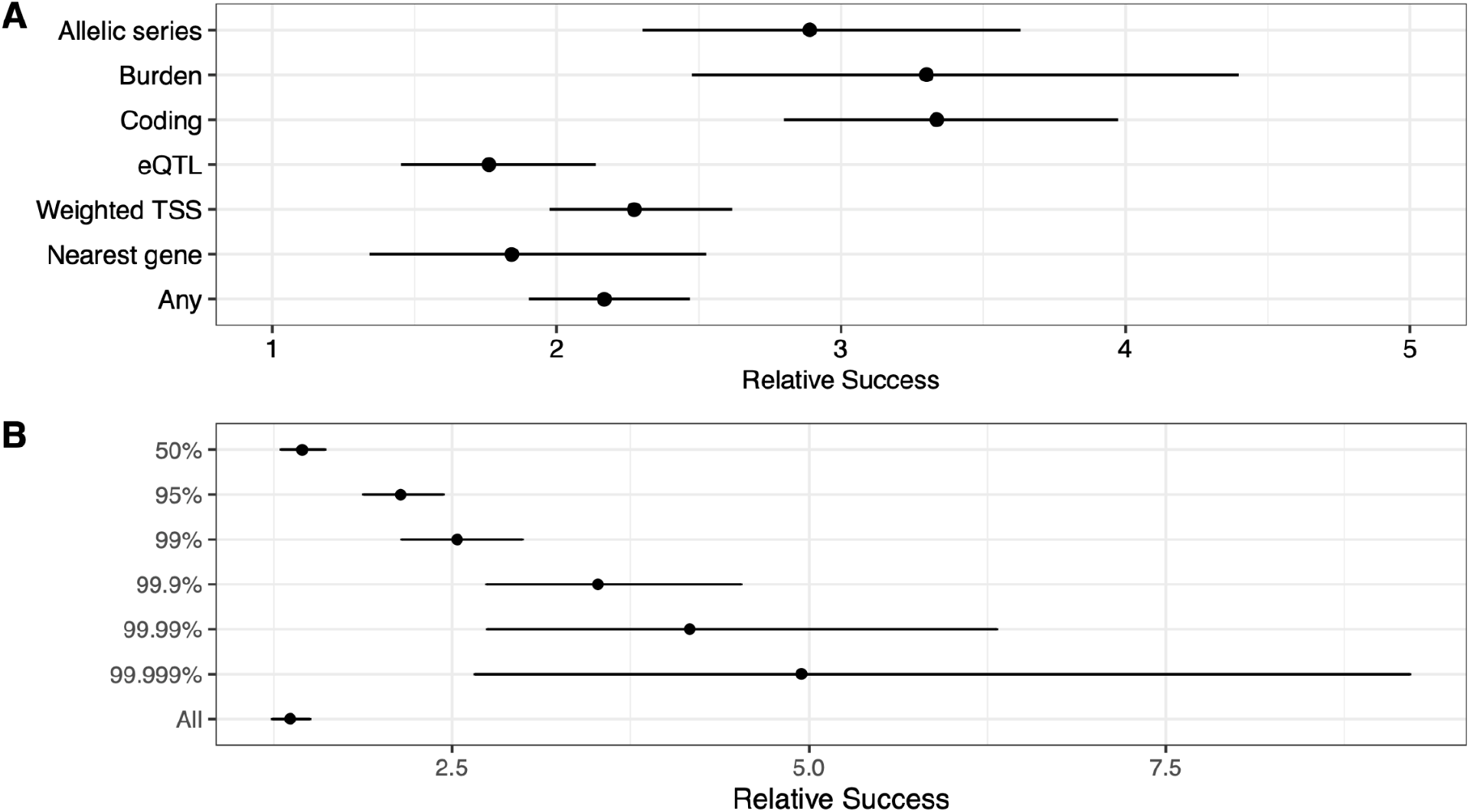
Relative success of clinical approval varies by gene-mapping criteria. A. Relative success for discrete V2G categories (see Methods); B. Relative success of clinical approval stratified by V2G score percentile in 23andMe data.

In order to systematically rank the strength of V2G hypotheses, we developed a V2G scoring framework leveraging the functional annotations with respect to a GWAS association. Using as truth set 1) the Open Targets Genetics gold standard dataset^36^ and 2) the evaluation dataset assembled leveraging fine-mapping results from UK Biobank^37^, we trained a machine learning model using per locus features on a gene-by-gene basis that includes: the number of QTL datasets, number of missense and loss-of-function variants, number of nearby genes, and Bayesian fine-mapping credible set weighted gene distance (see Methods for details) in a nested cross-validation framework (Supplementary Figure 4). The model achieves an overall ROCAUC of 98.2% and PRAUC of 92.7% (Supplementary Figure 5). We applied this model to the 23andMe European dataset and assigned V2G scores to 1.8 million gene-phenotype pairs across 181,279 loci. Within a single phenotype, the summation of scores across loci were assigned to each gene to capture independent V2G evidence and allelic series. Leveraging these scores in the Pharmaprojects framework, the relative clinical success for gene-phenotype pairs above the 99th, 99.9th, 99.99th, and 99.999th percentile V2G scores across all 1.8 million unique gene-phenotype pairs were 2.54, 3.52, 4.16, and 4.95 respectively (Figure 4B).

### Expansive set of target indication pairs yet to explore

To evaluate the opportunities for drug discovery and development from different sources of human genetic evidence, we assessed the number of supported T-I pairs that have been developed to at least phase I in Pharmaprojects and the total number of T-I pairs supported by human genetic evidence that have yet to be explored (Figure 5A). Considering those where the relative success is expected to be greater than 2.5, there are approximately 25,000 possible T-I pairs that have not been clinically pursued supported by 23andMe data and approximately 15,000 T-I pairs with support from Open Target Genetics (which includes data from GWAS Catalog, Neale UK Biobank, and FinnGen, etc), indicating great opportunities for future development. Among rare variants (MAF < 1%) where there is a 3-4x increased clinical success rate, we estimated over 2,000 T-I pairs supported by 23andMe data, and several hundred T-I pairs with support from the GWAS Catalog or the UK Biobank have not been pursued in the clinic (Figure 2C, D). As expected, we observed a negative correlation between RS and the number of tractable T-I pairs^40^, as fewer genes have high V2G scores and hence fewer T-I pairs developed or are open to pursuit. Regardless, thousands of T-I pairs with a predicted >3.5x relative success have not been developed clinically to date.

**Figure 5.**
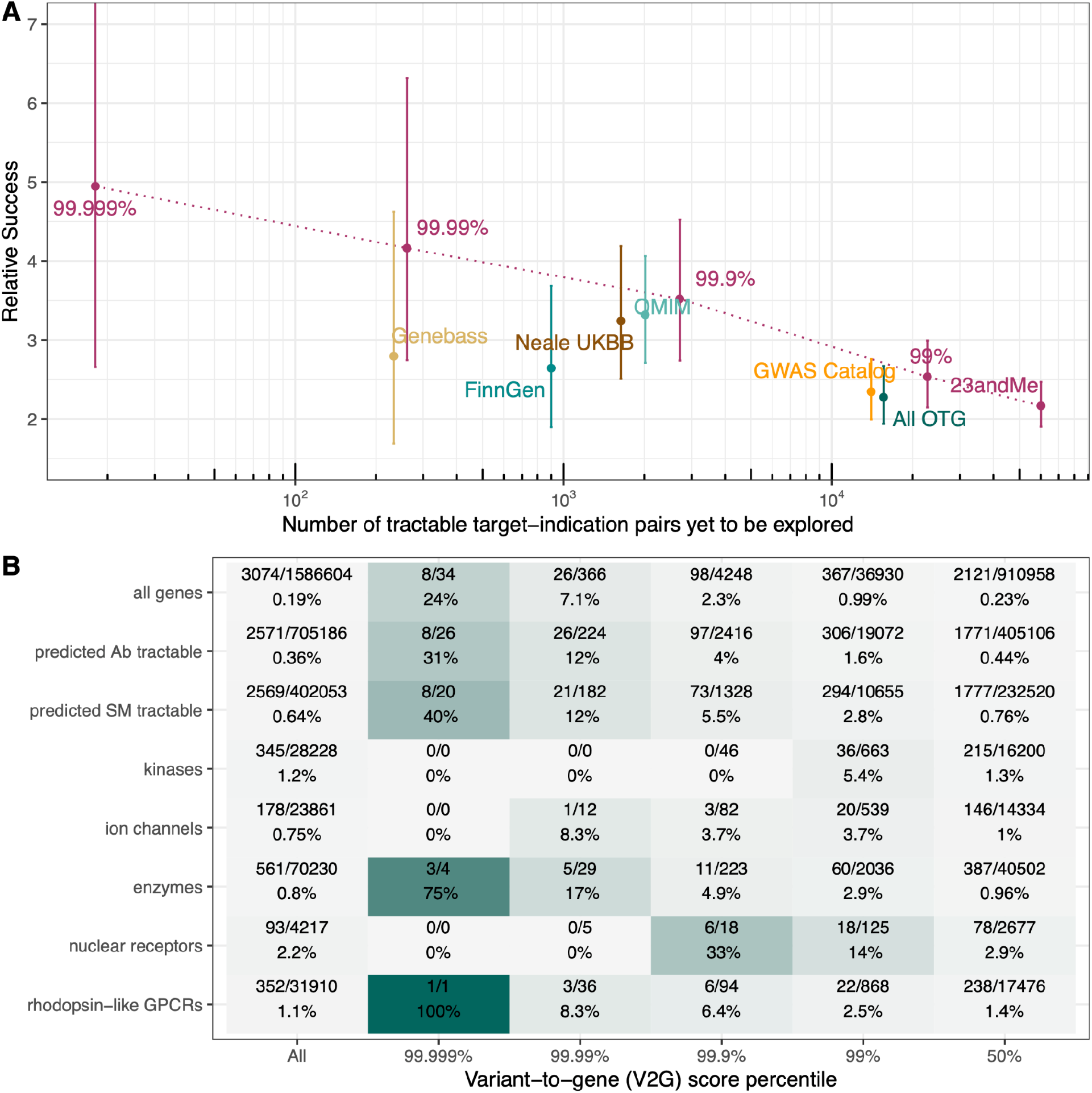
Expansive set of target indication pairs yet to explore by V2G criteria. A. Number of tractable T-I pairs relative to different genetic data sources, varying 23andMe V2G scoring criteria (points illustrate baseline V2G as in Figure 1, 99th, 99.9th, 99.99th, and 99.999th percentile of V2G scores across unique gene-phenotype pairs), dotted lines are linear interpolations from the point estimates, values for non-23andMe datasets were based on L2G mapping as in Minikel *et al*. 2024^23^; B. Proportion of T-I pairs supported by human-genetic evidence from 23andMe that have been pursued clinically to at least phase I by tractability and relevant protein classes, stratified by V2G score percentile.

Focusing on genetically supported T-I pairs that have been clinically pursued, we found that T-I pairs supported by greater V2G scores have much higher proportions that have been developed (Figure 5B). For example, 31% and 40% of tractable antibody and small-molecule T-I pairs in the 99.999th percentile V2G score have reached at least phase I, compared to 1.6% and 2.8% respectively for T-I pairs in the 99th percentile V2G score. This trend is consistent across a number of relevant protein classes, indicating greater V2G scores enrich for clinical success, and that genes in this category potentially have a large effect on disease. On the other hand, the T-I pairs that have been clinically assessed may represent the “low hanging fruit” in the explorable space of drug development. On average, across all genes, less than 1% T-I pairs have been evaluated clinically, and only 24% of gene-indication pairs in the 99.999th V2G score percentile have been considered. With rapid development of new modalities and innovation in all aspects of the drug development pipeline, we foresee exciting opportunities that lie ahead.

## Discussion

We leveraged 23andMe, the largest genotype-phenotype database in the world, to refine the estimate on enrichment of clinical success from human genetics supported programs. As is the case with clinically obtained phenotypes, programs with human genetics support derived from entirely self-reported phenotypes are 2-3 times more likely to succeed in the clinic, relative to the 10% industry average success rate. The improved power in discovering gene-trait associations by virtue of the large cohort size provides genetic support that can further increase the relative success for trial approvals. Many variants that are likely to have important functional consequences are often observed at lower frequencies in the general population. We demonstrated that very large cohorts with imputation of rare variants remains a highly effective approach to finding rare-variant associations in support of clinical programs, and programs with genetic support from rare-variant associations are 3-4 times more likely to be approved. Further, we confirm that gene mapping criteria can considerably influence the relative success of programs, and showcase that advanced gene scoring algorithms can improve the confidence in a program, resulting in up to 5-fold enrichment of clinical success.

One of the principal findings in this study is that genetic support from self-reported endpoints has similar impact on clinical success as endpoints derived from clinical sources (such as EMRs). Use of self-report through questionnaires and surveys is a highly scalable and efficient approach to phenotypic data collection, not limited to lifestyle and environment related phenotypes. For example, the latest multi-ancestry GWAS meta-analysis of Type II diabetes (T2D) largely drew cases from self-reported T2D individuals in the UKBB and the Icelandic cohort from deCode that also utilized self-reported diagnosis information^41^. While EMRs may contain more granular data, they may be fragmented due to the unique healthcare and insurance systems associated, and they are not exempt from misdiagnosis or changes in diagnosis and procedures for an individual over time. Self-reported data have its own difficulties, such as participants erroneously recalling diagnoses or misreporting of health-related information. Self-report may also be unsuitable for acquiring a broad range of lab values. Despite the GWAS catalog and a number of population cohorts and biobanks contain self-reported information and phenotypes derived from it, utilizing 23andMe data demonstrates that human genetics support from entirely self-reported phenotypes is equally enriched for clinical approvals, emphasizing the utility of leveraging self-reported data to maximize the statistical power of therapeutic discovery.

In contrast to conclusions from recent analysis^23^, we found that minor allele frequencies and effect sizes from GWAS influence the relative success estimates for program approvals, and that drug programs supported by rare and large effect associations have greater likelihood to be approved compared to common variant associations with small effects. This is in part due to the improved power to discover many rare-variant associations that provided genetic support for clinical programs from 23andMe, both with a large cohort size and the increased ability to impute rare variants at greater accuracies. However, there are a large number of low-frequency variants in the human population, rare-variant associations may be prone to false positives at the genome-wide significance p-value threshold (5e-8), and differences in LD patterns surrounding rare variants may pose additional challenges for gene mapping. Hence there remains important work to enable effective translation of rare-variant genetic associations to actionable clinical programs.

Whole exome and whole genome sequencing datasets can provide a great amount of value, especially for the discovery of rare variants that may influence gene regulation or function. However, the cost for sequencing hundreds of thousands to millions of individuals remains very high and unattainable for most study cohorts. Genotype imputation typically can not capture all variants that would be available in a sequencing dataset, however, leveraging the increasing availability of genome sequences for imputation into larger cohorts may significantly boost power for genetic association studies. Whole-exome and whole-genome sequences can not only offer confident genotype calls, but also identify a large number of coding variants that may have an impact on gene function. Given the continued improvement of phasing and imputation methods, it is now feasible to impute rare variants of very low frequencies, and hence cost-efficient to leverage sequences for imputation in a large cohort. In our V2G scoring model, coding variants have a higher relative influence compared to other features used (Supplementary figure 5), which may reflect the potential bias in the truth sets and that the 23andMe database is uniquely positioned to annotate a large number of loci with coding information to further strengthen V2G hypotheses. Accurate imputation of rare variants requires large panels of reference sequences, and critically, imputation quality remains lower in non-European cohorts^42^. This underscores the importance of continued efforts to enhance the size and representation in sequencing programs^43,44^.

We have focused primarily on genetic associations from the European population in this work. 23andMe has one of the largest non-European cohorts, with over 500,000 research participants with African American genetic ancestry for example. We see similar levels of relative success for clinical programs using data from ancestry groups that are not European (Supplementary figure 6), however, due to the large difference in discovery power, a majority of the genetically supported T-I pairs observed came from the European dataset. Regardless, we see a small number of supported T-I pairs that are only identified in non-European cohorts (Supplementary figure 7), and continued growth of these cohorts may present unique opportunities to identify genes important for disease biology.

In this work, we have treated the human genetics data sources as distinct (although the GWAS Catalog does contain some results from meta-analyses). However, in recent years, a number of initiatives have started to aggregate data across cohorts to both increase power and increase representation and diversity in the datasets, with a notable example being the Global Biobank Meta-analysis Initiative^45^. As we show in our work, greater statistical power in genetic associations provides further enrichment in success for the supported programs. This is likely due to a number of factors, including increased confidence in gene-mapping and decreased winner’s curse and other sources of false positives in the genetic findings. It is also worth noting that the T-I pairs supported by 23andMe only partially overlap with those in publicly available data sources (Figure 1, Supplementary figure 2), in aggregate providing support for ∼1500 T-I pairs in Pharmaprojects. This highlights the utility of combining data sources each measuring some discrete entities, and in aggregate can not only further improve discovery power but also provide a broad coverage of programs that are more likely to progress.

In conclusion, we leveraged the 23andMe cohort to explore how both the scale of the human genetics data and improved variant to gene mapping from genetic associations influence estimates of relative success for clinical approvals of therapeutic programs. We found that programs with genetic support from self-reported data, just as for clinically derived data, are 2-3 times likely to succeed in the clinic. Genetic support from rare, large effect associations have increased likelihood of success, and improved statistical power as well as improved gene mapping can further enhance the relative success rate for trials.

## Acknowledgements

We would like to thank David A. Hinds, Michael V. Holmes, and Pierre Fontanillas for discussion and critical feedback. We would like to thank the research participants and employees of 23andMe for making this work possible. The following members of the 23andMe Research Team contributed to this study: Stella Aslibekyan, Adam Auton, Elizabeth Babalola, Robert K. Bell, Jessica Bielenberg, Ninad S. Chaudhary, Zayn Cochinwala, Sayantan Das, Emily DelloRusso, Payam Dibaeinia, Sarah L. Elson, Nicholas Eriksson, Chris Eijsbouts, Teresa Filshtein, Pierre Fontanillas, Davide Foletti, Will Freyman, Zach Fuller, Julie M. Granka, Chris German, Éadaoin Harney, Alejandro Hernandez, Barry Hicks, David A. Hinds, M. Reza Jabalameli, Ethan M. Jewett, Yunxuan Jiang, Sotiris Karagounis, Lucy Kaufmann, Matt Kmiecik, Katelyn Kukar, Alan Kwong, Keng-Han Lin, Yanyu Liang, Bianca A. Llamas, Aly Khan, Steven J. Micheletti, Matthew H. McIntyre, Meghan E. Moreno, Priyanka Nandakumar, Dominique T. Nguyen, Jared O’Connell, Steven J. Pitts, G. David Poznik, Alexandra Reynoso, Shubham Saini, Morgan Schumacher, Leah Selcer, Anjali J. Shastri, Jingchunzi Shi, Suyash Shringarpure, Keaton Stagaman, Teague Sterling, Qiaojuan Jane Su, Joyce Y. Tung, Susana A. Tat, Vinh Tran, Xin Wang, Wei Wang, Catherine H. Weldon, Amy L. Williams, Peter Wilton.

## Competing Interests

X.W., S.K., S.S.S., R.S., Q.J.S, V.V., S.J.P., A.A. are employed by and hold stock or stock options in 23andMe, Inc.

## Data availability statement

Data sharing pertaining to 23andMe data supporting the findings of this study may be made available upon reasonable request or collaboration. Non-23andMe data are publicly available. Download links: OpenTargets - https://github.com/opentargets/genetics-gold-standards/, Pharmaprojects and benchmarking datasets - https://github.com/ericminikel/genetic_support, Weeks et al. - https://www.finucanelab.org/data.

## Notes

### Funding Statement

This study did not receive any funding

### Author Declarations

Participants provided informed consent and volunteered to participate in the research online, under a protocol approved by the external AAHRPP-accredited IRB, Ethical & Independent (E&I) Review Services. As of 2022, E&I Review Services is part of Salus IRB (https://www.versiticlinicaltrials.org/salusirb). All research was performed in accordance with relevant guidelines/regulations, including the Declaration of Helsinki.

