## Supplemental Figures for "The impact on clinical success from the 23andMe cohort"

### Supplementary materials

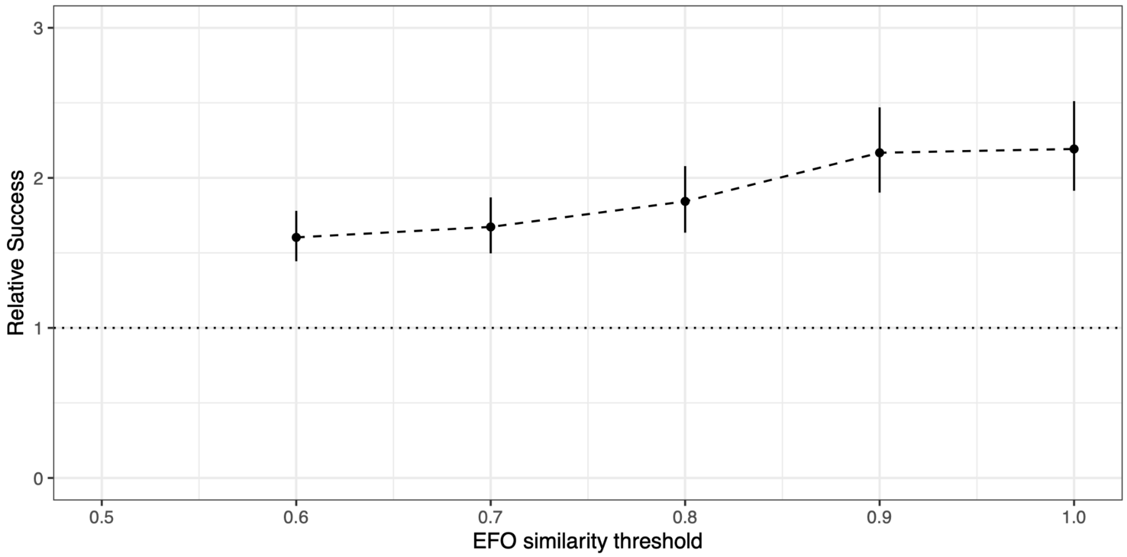


**Supplementary Figure 1. Sensitivity of relative success in 23andMe data to EFO similarity threshold.** EFO similarity threshold used for considering a 23andMe phenotype a “match” to Pharmaprojects indications were varied from 0.6 to 1.0, and relative success of clinical approval were calculated for each threshold.

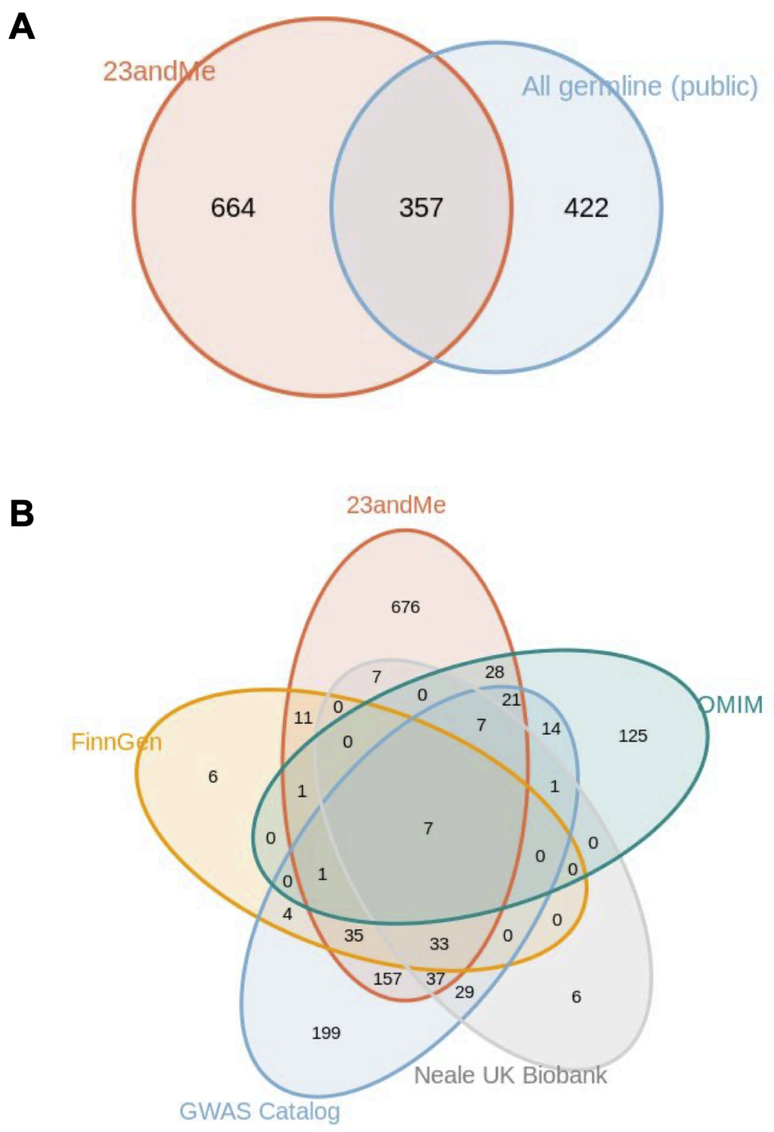


**Supplementary Figure 2. Overlap of genetically supported target-indication pairs across five datasets**. Genetically supported target-indication pairs in Pharmaprojects that have reached at least phase I A. from 23andMe versus all public germline data (including data from the GWAS Catalog, UK Biobank, FinnGen, OMIM, PICCOLO, and genebass.org)[1]; B. from 23andMe, the GWAS Catalog, UK Biobank, FinnGen or OMIM and whether they are shared across two or more datasets.

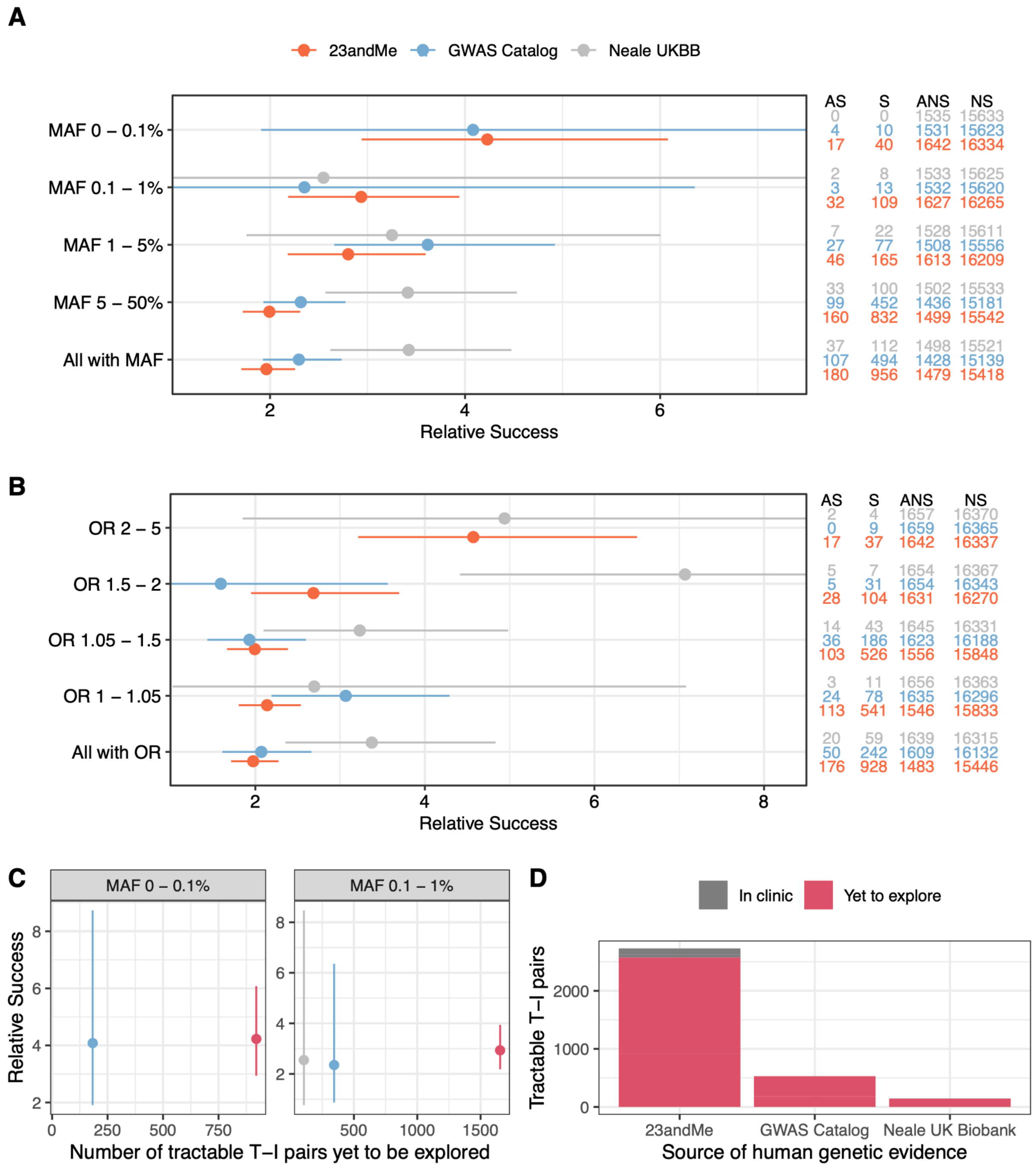


**Supplementary Figure 3. Relative success of rare, large effect genetic associations and clinical opportunities removing OMIM.** A-B. Relative success in 23andMe data stratified by minor allele frequency (MAF) and odds ratio (OR) in comparison to the GWAS Catalog and the UK Biobank; C. The number of tractable target-indication (T-I) pairs yet to be explored, stratified by MAF among variants with MAF < 1%; D. Comparison of tractable T-I pairs already in clinic versus those yet to be explored by source of human genetic evidence (MAF < 1%). All T-I pairs that are identified via OMIM were removed.

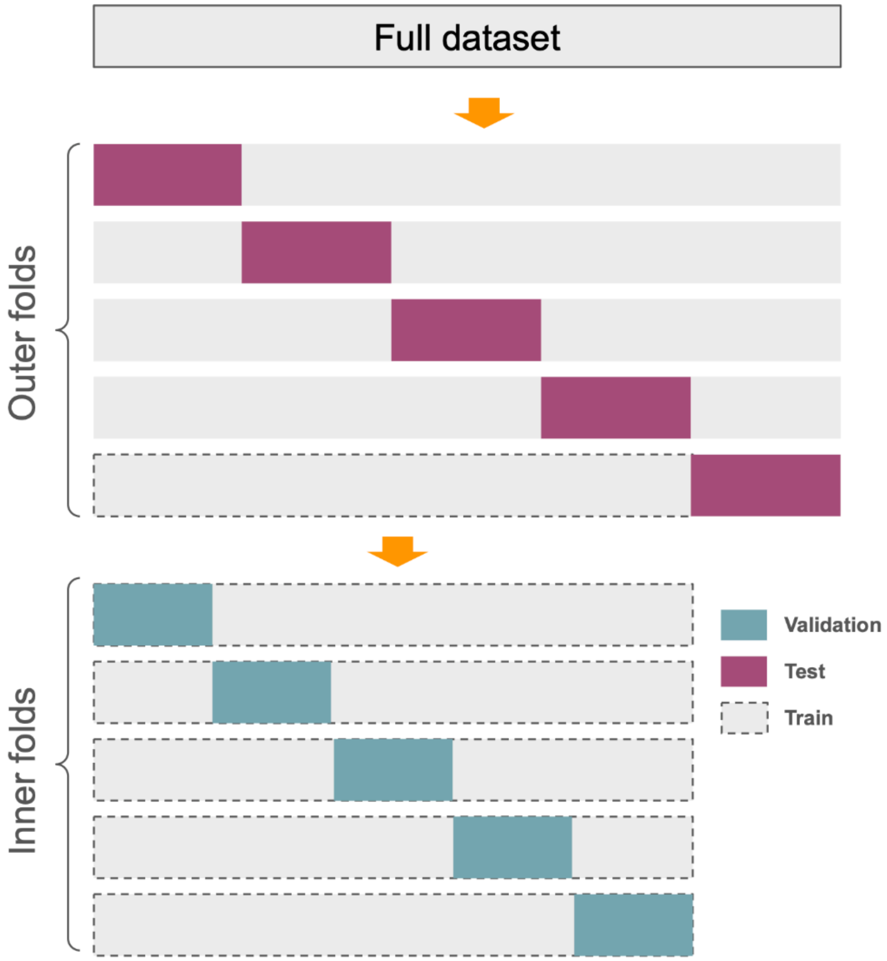


**Supplementary Figure 4. Schematic of nested cross-validation framework.** Outer folds were segmented by chromosomes such that each chromosome belonged to one fold and one fold only. For each outer fold, five inner cross-validation folds were used. The test set is hence entirely independent from the training and validation set used in model building and parameter tuning.

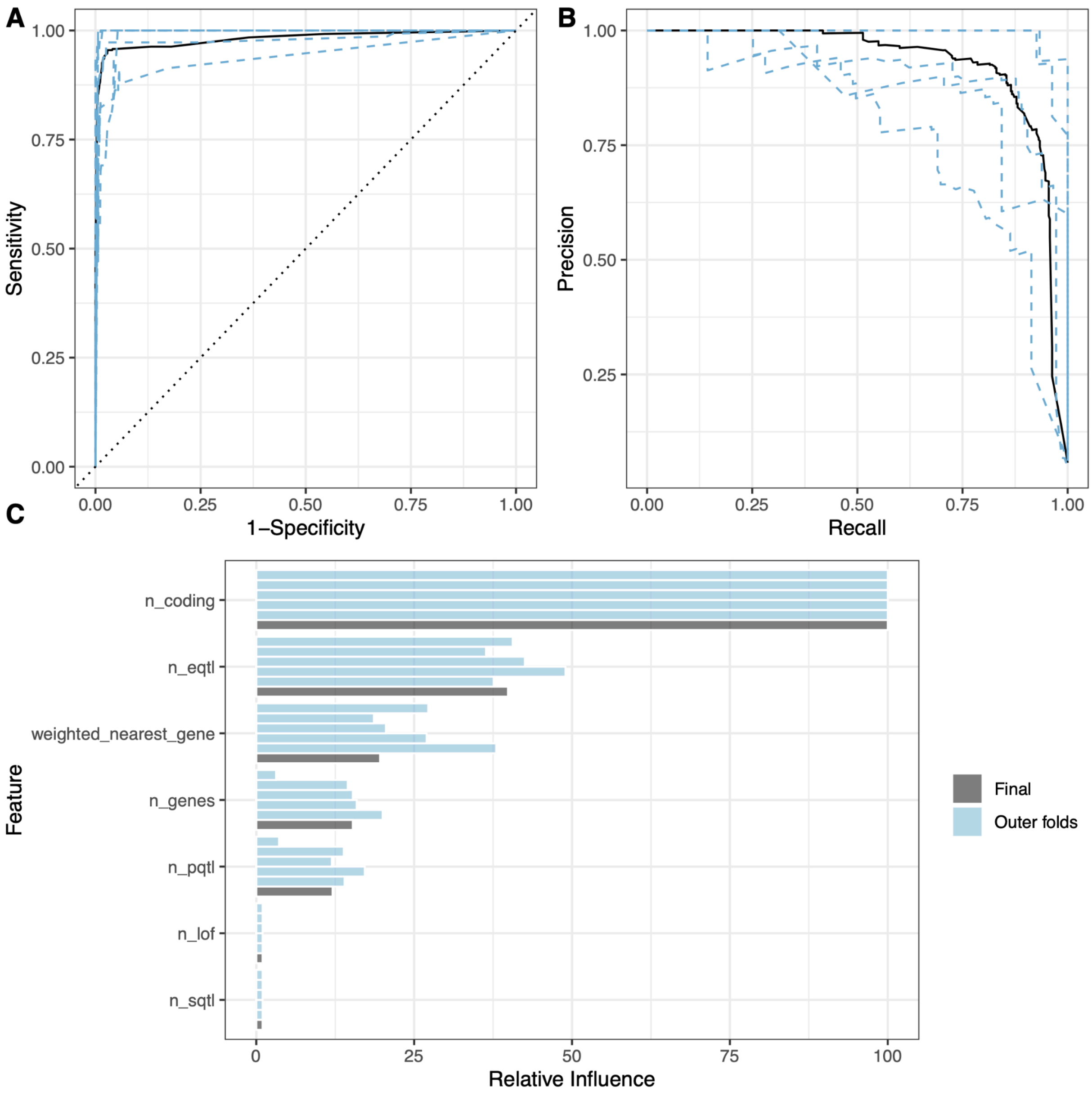


**Supplementary Figure 5. Performance of V2G scoring model.** A. ROC performance curve. Lighter colored curves show ROC performance per outer CV fold; B. PR performance curve. Lighter colored curves show PR performance per outer CV fold; C. Relative importance for each feature used in the model, as described in Friedman (2001)[2] and implemented in the *gbm* package in R.

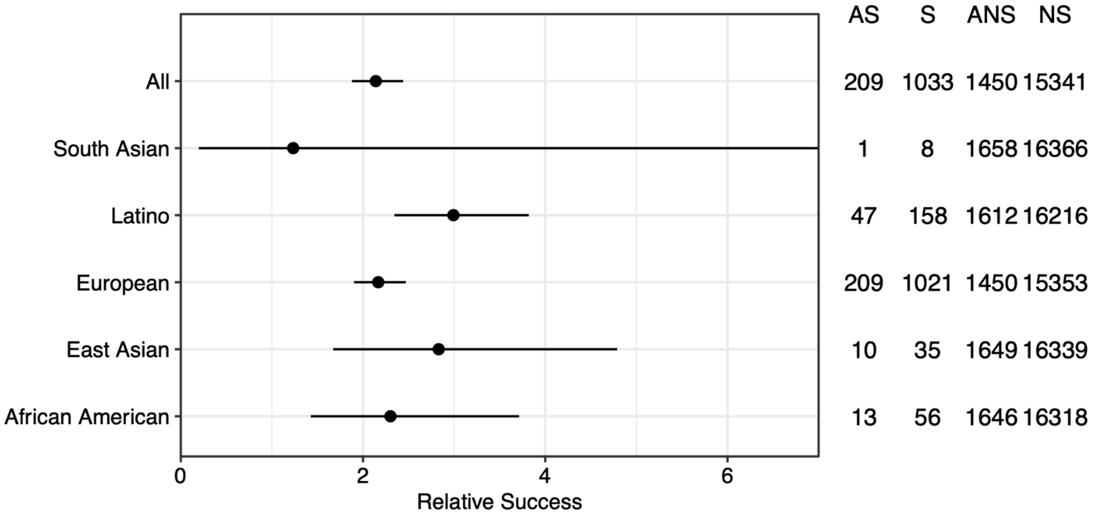


**Supplementary Figure 6. Relative success stratified by genetic ancestry groups.** Genetically supported target-indication pairs were identified for each of five genetic ancestry groups (African American, East Asian, European, Latino, South Asian), and relative success was estimated per group (see Methods). The overall relative success was estimated by combining all groups (All).

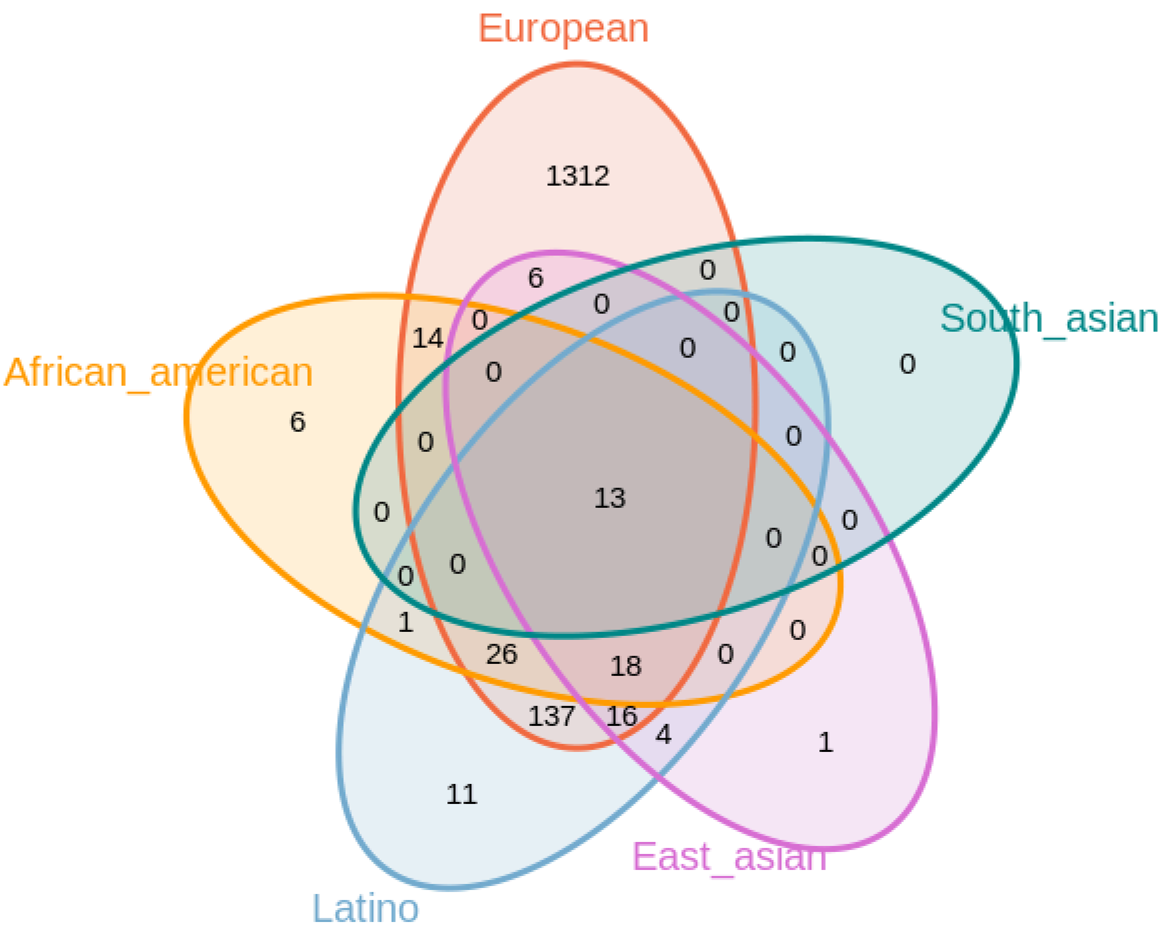


**Supplementary Figure 7. Overlap of genetically supported target-indication pairs across genetic ancestry groups.** Genetically supported target-indication pairs were identified for each of five genetic ancestry groups (African American, East Asian, European, Latino, South Asian), their overlaps are shown in the Venn diagram.

**Supplementary Table 1. Mappings between Pharmaprojects indications and 23andMe phenotypes.** Ontology mappings associated with indications in Pharmaprojects (indication_mesh_id, indication_mesh_term, indication_efo_id) were mapped to 23andMe phenotypes (assoc_efo_id, assoc_efo_term, phenotype) at the specified similarity threshold (sim). Details see methods.
